# Increased Influenza Severity in Children in the Wake of SARS-CoV-2

**DOI:** 10.1101/2023.03.11.23286858

**Authors:** Gregory Hoy, Hannah E. Maier, Guillermina Kuan, Nery Sánchez, Roger López, Alyssa Meyers, Miguel Plazaola, Sergio Ojeda, Angel Balmaseda, Aubree Gordon

**Affiliations:** Department of Epidemiology, School of Public Health, University of Michigan, Ann Arbor, Michigan, USA; Sustainable Sciences Institute, Managua, Nicaragua; Centro de Salud Sócrates Flores Vivas, Ministry of Health, Managua, Nicaragua; Laboratorio Nacional de Virología, Centro Nacional de Diagnóstico y Referencia, Ministry of Health, Managua, Nicaragua

**Author notes:** Correspondence: Aubree Gordon, PhD, 5622 SPH I, School of Public Health, 1415 Washington Heights, Ann Arbor, MI, USA 48109-2029 e. Alternative contact: Gregory Hoy, 1415 Washington Heights, Ann Arbor, MI, USA 48109-2029, e.

**Keywords:** influenza, pediatrics, influenza severity, SARS-CoV-2, global health

## Abstract

The SARS-CoV-2 pandemic and subsequent interruption of influenza circulation has lowered population immunity to influenza, especially among children with few pre-pandemic exposures. We compared the incidence and severity of influenza A/H3N2 and influenza B/Victoria between 2022 and two pre-pandemic seasons and found an increased frequency of severe influenza in 2022.

---

In the two years following the emergence of SARS-CoV-2, global circulation of influenza was low [1]. This gap in influenza circulation, along with decreased uptake of influenza vaccination, has lowered population immunity to influenza, which could lead to increased severity of influenza epidemics [2,3]. Children experience symptomatic influenza infections more frequently than adults and bear a disproportionate portion of the burden of influenza[4,5]. In addition, children have fewer total lifetime influenza exposures and are normally exposed to influenza in the first few years of life, so they may be at even greater risk of severe influenza during rebound seasons due to fewer prior exposures and changes in the age patterns of first influenza exposure[6].

In 2022, influenza began to re-circulate in much of the world with sporadic seasonality, limited antigenic diversity of circulating strains, and the absence of B/Yamagata circulation globally [2,7]. However, the severity of influenza in children in 2022 has not been compared to prior seasons, and the clinical impact of waning influenza immunity during the gap in influenza circulation has not been described. Using data from a prospective cohort study of children in Managua, Nicaragua, we describe the incidence and severity of influenza in 2022 and examine whether the incidence and severity of symptomatic influenza infections is greater in 2022 compared to several pre-pandemic seasons.

## Methods

The Nicaraguan Pediatric Influenza Cohort Study (NPICS) is a prospective pediatric cohort study based in Managua, Nicaragua that enrolls healthy children aged 0-14 [8]. If enrolled children become ill, they are examined by study physicians and tested for influenza if they have pneumonia or severe respiratory disease, or if they meet age-based symptom criteria (Supplemental Table 1). Combined nasal/oropharyngeal samples were obtained and tested for influenza using real-time reverse-transcription polymerase chain reaction (RT-PCR) using validated CDC protocols[9]. If positive for influenza, subtype or lineage was obtained following CDC protocols [10].

Both influenza A/H3N2 and B/Victoria circulated in the NPICS in 2022; thus, the two most recent pre-2020 A/H3N2 seasons (2017-2018, 2019-2020) and B/Victoria seasons (2017-2018, 2018-2019) were selected for comparison[11]. Clinical symptom and diagnosis data were obtained from health center visits up to 10 days before and 30 days after the date of influenza RT-PCR positivity. Severe infection was defined as an influenza infection associated with hospitalization or clinically diagnosed pneumonia. Frequencies of symptoms and severe cases were compared using χ2 or Fisher exact tests. Analyses were performed using SAS 9.4 and R version 4.2.2. This study was approved by the institutional review boards at the Nicaraguan Ministry of Health and the University of Michigan.

## Results

### A/H3N2

A total of 330 A/H3N2 cases occurred in NPICS in the 2017-2018 and 2019-2020 influenza seasons, and 198 A/H3N2 cases occurred in the 2022 influenza season (Figure 1a). No A/H3N2 infections were detected in the cohort between February 2020 and January 2022. Children infected with A/H3N2 in 2022 were older on average than those infected with A/H3N2 in the pre-2020 seasons, with an average age of infection in the 2022 season of 6.5 years, compared to 5.2 years for the pre-2020 seasons (p = 0.008, Figure 1b). The incidence of A/H3N2 per 100 person-years was significantly higher in 2022, with incidence estimates of 9.15 (7.83-10.69) in 2017-2018, 11.1 (9.61-12.76) in 2019-2020, and 15.8 (14.00-17.83) in 2022. When stratified by age, incidence in 2022 was higher in children aged 10-14, with no difference in incidence in children aged 0-4 (Figure 1c). Vaccination rates in the cohort were low overall, and children were more likely to have been vaccinated for influenza in 2022 compared to pre-2020 seasons; 19 (9.6%) children were vaccinated in the 6 months prior to infection in 2022, compared to 15 (4.5%) children in the pre-2022 seasons (p = 0.023). The symptomatic presentation of 2022 cases was similar to pre-2020 cases, with no differences in the frequency of subjective fever, cough, gastrointestinal (GI) symptoms, or Influenza-like-illness (ILI); however, objective fever, sore throat and headache were less frequently reported in 2022 cases than pre-2020 cases (Supplemental Table 2). Children aged 0-14 were nearly 3 times more likely to experience a severe case of A/H3N2 in 2022 compared to the pre-2020 seasons. Of the 198 cases in 2022, 7 (3.5%) of them were severe; of the 330 cases in the pre-2020 seasons, 4 (1.2%) of them were severe (p = 0.111) (Figure 1d, Supplemental Table 3). Influenza cases were significantly more severe in children aged 0-4 in 2022; 6 of 87 (6.9%) cases were severe in 2022, compared to 3 of 189 (1.6%) cases in the pre-2020 seasons (p = 0.030; Figure 1d).

**Figure 1.**
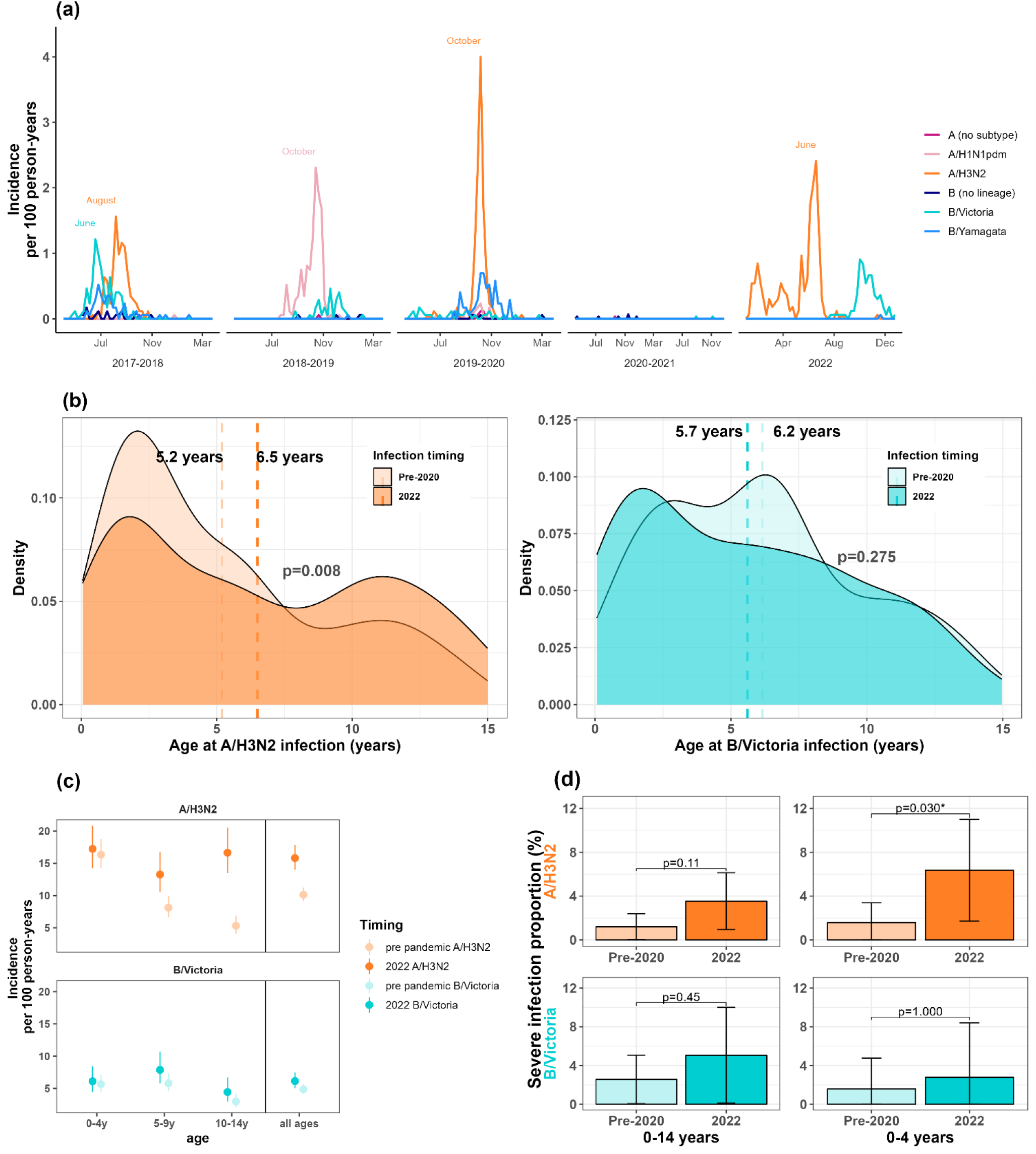
(a) Influenza incidence rate in the Nicaraguan Pediatric Influenza Cohort Study, by season. (b) Age distribution of A/H3N2 and B/Victoria cases, pre-2020 vs. 2022 (Wilcoxon Rank Sum Test). (c) Comparison of influenza incidence rate pre-2020 vs. 2022, by age. (d) Frequency of severe influenza infection, pre-2020 vs. 2022 (Fisher’s Exact Test).

### B/Victoria

A total of 156 B/Victoria cases occurred in NPICS in the 2017-2018 and 2018-2019 seasons, and 80 cases occurred in the 2022 influenza season (Figure 1a). Only three infections were detected in the cohort between February 2020 and July 2022. There was no difference in the age distribution of B/Victoria cases in 2022 compared to pre-2020 seasons; the average age of infection in the 2022 season was 5.7 years, compared to 6.2 years in the pre-2020 seasons (p = 0.275, Figure 1b).The incidence of B/Victoria per 100 person-years was 7.29 (6.13-8.69) for 2017-2018, 2.48 (1.84-3.35) for 2018-2019, and 6.15 (5.07-7.47) for 2022 (Figure 1c). Influenza B/Victoria cases in 2022 were also more likely to have been vaccinated for influenza in the 6 months prior to infection (18.8% vs. 0.6%, p < 0.001). Compared to pre-2020 cases, 2022 cases had a higher frequency of many signs and symptoms, including objective fever, cough, and headache (Supplemental Table 4). Influenza B/Victoria cases were not more severe than cases in the pre-2020 (p = 0.447; Figure 1d, Supplemental Table 5).

## Discussion

Low circulation of influenza during the first two years of the SARS-CoV-2 pandemic was predicted to increase the incidence and clinical severity of influenza infections during rebound influenza epidemics due to decreased population immunity. We found that children infected with A/H3N2 in 2022 were a year older, on average than children infected with A/H3N2 in the pre-2020 seasons, possibly representing a delay in the timing of the first few lifetime influenza infections. Though this age pattern is unlikely to become entrenched without another multi-year gap in influenza circulation, it suggests that immunologic age as a function of exposure history may matter more than biologic age for the risk of influenza infections in young children.

Older children normally have some degree of immunity to influenza that is continually boosted by repeated childhood infections and seasonal circulation; we hypothesized that this immunity waned while influenza was not circulating and predicted we would see increased symptomatic influenza incidence in older children once influenza returned. We found an increase in A/H3N2 incidence in children aged 10-14, but not in children aged 0-4. The similar rate of influenza infection in young children between seasons suggests that the overall force of infection was likely similar, and the increase observed in older children supports the notion that waning influenza immunity due to SARS-CoV-2 may have increased either the underlying susceptibility to infection and/or the proportion of infections that were symptomatic and therefore detected.

In addition to an increase in susceptibility to symptomatic influenza infection in older children, we also observed higher severity of influenza cases. Children in the cohort in 2022 were about three times more likely to experience a severe infection. Children under 4 years of age showed a significant increase in the proportion of severe A/H3N2 infections in 2022. This increased severity may be partially attributable to waning immunity in these children due to decreased influenza circulation during 2020 and 2021, as well as other factors, such as decreased passive immunity in the first 6 months of life due to decreased maternal influenza infection and vaccination during periods of low influenza circulation. Influenza A/H3N2 infections were also less likely to experience symptoms related to a strong immune response to infection such as objective fever, sore throat, and headache, which also supports the notion of lower pre-existing immunity in children in 2022[12].

Unlike with A/H3N2, the age distribution and incidence of B/Victoria cases in 2022 was not significantly different than the pre-2020 seasons, likely because 2-year gaps in influenza B circulation occur commonly in our setting [11]. Severe B/Victoria was more likely to occur in 2022 than in the pre-2020 B/Victoria seasons; however, one limitation of our study it that the small sample limits the statistical power of these comparisons.

## Conclusion

Using data from one of the largest and longest-running prospective pediatric cohorts of influenza, we found that 2022 A/H3N2 infections had higher incidence in older children, and 2022 infections were 3-4 times as likely to be severe compared to pre-2020 A/H3N2 infections. This increased severity may represent waning influenza immunity in children due to the SARS-CoV-2 pandemic.

## Data Availability

All data produced in the present study are available upon reasonable request to the authors.

## ACKNOWLEDGEMENTS

We thank the study participants and the many dedicated study personnel in Nicaragua at the Centro Nacional de Diagnóstico y Referencia and the Sócrates Flores Vivas Health Center.

