## Supplemental Materials for "Increased Influenza Severity in Children in the Wake of SARS-CoV-2"

| Children < 2 | Children ≥2 |
| --- | --- |
| Illness onset ≤4 days prior  AND  Fever or feverishness | Illness onset ≤4 days prior  **AND**  Fever or feverishness  **AND**  Cough **OR** sore throat **OR** rhinorrhea |

**Supplemental Table 1. Influenza testing criteria in NPICS**

**Supplemental Table 2. A/H3N2 symptom comparison, pre-2020 vs. 2022**

|  | Pre-2020 (n=330) | 2022  (n=198) | p-value |
| --- | --- | --- | --- |
| Symptom - n (%) |  |  |  |
| Subjective fever | 324 (98.2) | 198 (100.0) | 0.088 |
| Temp. ≥37.8^◦^C | 225 (68.2) | 115 (58.1) | 0.024* |
| Cough | 291 (88.2) | 170 (85.9) | 0.500 |
| Rhinorrhea | 306 (92.7) | 169 (85.4) | 0.010* |
| Congestion | 136 (41.2) | 82 (41.4) | 1.000 |
| Sore throat | 105 (31.8) | 39 (19.7) | 0.003* |
| Headache | 49 (14.9) | 13 (6.6) | 0.005* |
| Any GI symptom | 63 (19.1) | 35 (17.7) | 0.730 |
| Vomiting | 27 (8.2) | 18 (9.1) | 0.749 |
| Nausea | 6 (1.8) | 1 (0.5) | 0.265 |
| Diarrhea | 25 (7.6) | 24 (12.1) | 0.090 |
| Abdominal pain | 17 (5.2) | 5 (2.5) | 0.179 |
| Influenza-like illness | 147 (44.6) | 83 (41.9) | 0.587 |

p-values from Fisher’s exact tests. Significant p-values at alpha = 0.05 indicated by *.

|  | Pre-2020 (n=330) | 2022  (n=198) | p-value |
| --- | --- | --- | --- |
| Severe illness – n (%) |  |  |  |
| A/H3N2 |  |  |  |
| Overall | 4 (1.2) | 7 (3.5) | 0.111 |
| 0-4 years | 3 (1.6) | 6 (6.9) | 0.030* |
| 5-14 years | 1 (0.7) | 1 (0.9) | 1.000 |

**Supplemental Table 3. A/H3N2 severe illness, by age group**

p-values from Fisher’s exact tests. Significant p-values at alpha = 0.05 indicated by *.

**Supplemental Table 4. B/Victoria symptom comparison, pre-2020 vs. 2022**

|  | Pre-2020 (n=156) | 2022  (n=80) | p-value |
| --- | --- | --- | --- |
| Symptom - n (%) |  |  |  |
| Subjective fever | 151 (96.8) | 80 (100.0) | 0.170 |
| Temp. ≥37.8^◦^C | 92 (59.0) | 61 (76.3) | 0.010* |
| Cough | 134 (85.9) | 59 (73.8) | 0.032* |
| Rhinorrhea | 118 (75.6) | 69 (86.3) | 0.063* |
| Congestion | 52 (33.3) | 39 (48.7) | 0.024* |
| Sore throat | 59 (37.8) | 27 (33.7) | 0.570 |
| Headache | 20 (12.8) | 24 (30.0) | 0.002* |
| Any GI symptom | 23 (14.7) | 16 (20.0) | 0.355 |
| Vomiting | 7 (4.5) | 11 (13.8) | 0.018* |
| Nausea | 2 (1.28) | 3 (3.8) | 0.340 |
| Diarrhea | 14 (9.0) | 5 (6.3) | 0.615 |
| Abdominal pain | 8 (5.1) | 4 (5.0) | 1.000 |
| Influenza-like illness | 57 (36.5) | 39 (48.8) | 0.097 |

p-values from Fisher’s exact tests. Significant p-values at alpha = 0.05 indicated by *.

**Supplemental Table 5. B/Victoria severe illness, by age group**

p-values from Fisher’s exact tests. Significant p-values at alpha = 0.05 indicated by *.

|  | Pre-2020 (n=156) | 2022  (n=80) | p-value |
| --- | --- | --- | --- |
| Severe illness – n (%) |  |  |  |
| B/Victoria |  |  |  |
| Overall | 4 (2.6) | 5 (6.3) | 0.172 |
| 0-4 years | 1 (1.6) | 1 (2.8) | 1.000 |
| 5-14 years | 3 (3.2) | 4 (9.1) | 0.211 |
